# Family history and polygenic risk of cardiovascular disease: independent factors associated with secondary cardiovascular events in patients undergoing carotid endarterectomy

**DOI:** 10.1101/19006718

**Authors:** Nathalie Timmerman, Dominique P.V. de Kleijn, Gert J. de Borst, Hester M. den Ruijter, Folkert W. Asselbergs, Gerard Pasterkamp, Saskia Haitjema, Sander W. van der Laan

## Abstract

**Background:** Family history (FHx) of cardiovascular disease (CVD) is a risk factor for CVD and a proxy for cardiovascular heritability. Polygenic risk scores (PRS) summarizing >1 million variants for coronary artery disease (CAD) are associated with incident and recurrent CAD events. However, little is known about the influence of FHx or PRS on secondary cardiovascular events (sCVE) in patients undergoing carotid endarterectomy (CEA).

**Methods:** We included 1,788 CEA patients from the Athero-Express Biobank. A weighted PRS for CAD including 1.7 million variants was calculated (MetaGRS). The composite endpoint of sCVE during three years follow-up included coronary, cerebrovascular and peripheral events and cardiovascular death. We assessed the impact of FHx and MetaGRS on sCVE and carotid plaque composition.

**Results:** Positive FHx was associated with a higher 3-year risk of sCVE independent of cardiovascular risk factors and MetaGRS (adjusted HR 1.40, 95%CI 1.07-1.82, p=0.013). Patients in the highest MetaGRS quintile had a higher 3-year risk of sCVE compared to the rest of the cohort independent of cardiovascular risk factors including FHx (adjusted HR 1.35, 95%CI 1.01-1.79, p=0.043), and their atherosclerotic plaques contained more fat (adjusted OR 1.59, 95%CI, 1.11-2.29, p=0.013) and more macrophages (OR 1.49, 95%CI 1.12-1.99, p=0.006).

**Conclusion:** In CEA patients, both positive FHx and higher MetaGRS were independently associated with increased risk of sCVE. Moreover, higher MetaGRS was associated with vulnerable plaque characteristics. Future studies should unravel underlying mechanisms and focus on the added value of PRS and FHx in individual risk prediction for sCVE.

## Introduction

Family history of cardiovascular disease (FHx) is a major risk factor for primary cardiovascular disease (CVD) and serves as a surrogate for genetic predisposition.(1, 2) Risk prediction for secondary cardiovascular events remains challenging as traditional risk factors have limited discriminative performance.(3) The main underlying mechanism of CVD is atherosclerosis, and atherosclerotic plaque composition, exemplified by intraplaque haemorrhage (IPH), was shown to be associated to adverse secondary cardiovascular events (sCVE).(4) Yet, the relevance of FHx for secondary outcome of cardiovascular events is still unclear.(5–9)

Large-scale genome-wide association studies (GWAS) have identified hundreds of common genetic variants (single-nucleotide polymorphisms or SNPs) robustly associated with coronary artery disease (CAD)(10–14) and ischemic stroke(15–17) predisposition, albeit with small individual effects. Exact pathobiological mechanisms leading to cardiovascular symptoms are still poorly understood, but CAD- and ischemic stroke genetic variants were previously associated to atherosclerotic plaque composition.(18)

Polygenic risk scores (PRS) summarize the small individual genetic effects into a quantitative measure of genetic disease susceptibility. Recent studies showed that PRS were strongly correlated with prevalent and incident CAD independent of traditional risk factors including family history in the UK Biobank population.(19, 20) For example, individuals with higher scores of the MetaGRS (a PRS for CAD including 1.7 million SNPs) were at 1.7-4.2 fold higher risk for a first coronary event compared to individuals with lower MetaGRS scores. In addition, a very recent study in a French-Canadian population with established CAD showed that MetaGRS was also associated with increased risk of recurrent CAD events.(21)

As atherosclerosis is known for its complex nature with genetic overlap in different diseases such as stroke, CAD, abdominal aortic aneurysm (AAA) and peripheral artery disease (PAD)(22), we aimed to investigate the association between MetaGRS and long-term secondary cardiovascular events in a different cohort of severe atherosclerotic patients with carotid artery stenosis undergoing carotid endarterectomy. Given that FHx is used in clinical practice as a derivative of genetic background, we also examined the association between FHx and secondary cardiovascular events. Moreover, to explore possible underlying pathophysiological mechanisms, we studied the impact of MetaGRS and FHx on carotid histological plaque characteristics.

## Methods

### Athero-Express Biobank

All patients in this study were included in the Athero-Express Biobank, a prospective cohort study that included consecutive patients with severe carotid artery stenosis undergoing CEA in two large tertiary referral hospitals in The Netherlands, the University Medical Centre Utrecht (inclusion is ongoing) and the St. Antonius Hospital Nieuwegein (inclusion until 2014).(23) The study design has been published before.(23) In short, patient characteristics, such as demographics, cardiovascular risk factors including medical history, medication use, and FHx for cardiovascular disease, were obtained through standardized questionnaires and checked in electronic health records. Preoperative blood samples were drawn. The atherosclerotic plaque obtained during surgery was collected and immunohistochemically analysed for plaque characteristics. Patients were followed up for three years after surgery for the occurrence of secondary cardiovascular events through standardized questionnaires and by checking electronic health records. General practitioners were consulted in case of no response to questionnaires or in order to obtain further information regarding reported cardiovascular events. Patients operated for restenosis (6% of 2,044 eligible patients for this study) were excluded because these differ in future cardiovascular event risk.(24) Thus for the current study, a total of 1,788 patients operated from March 2002 until July 2016 had available 3-year follow-up data and FHx data, and were included for analysis. Of these, 1,551/1,788 (87%) patients had available histological carotid plaque data. A total of 1,319/1,788 (74%) patients had available genotype data of whom 1,301 (98%) also had histological carotid plaque data. This study was performed according to the Declaration of Helsinki and was approved by the local ethics committee of both hospitals. Patients provided written informed consent before study participation.

### Definitions

A positive FHx was defined as having a first-degree relative (either a parent or sibling) with onset of cardiovascular disease (myocardial infarction (MI), coronary artery stenosis, stroke, AAA, or cardiovascular death including sudden death) before the age of 60 years. The primary outcome of this study was defined as a composite secondary cardiovascular event (sCVE) within three years of follow-up including fatal or non-fatal MI, fatal or non-fatal stroke, ruptured AAA, fatal cardiac failure, coronary or peripheral interventions (either percutaneous or bypass surgery), leg amputation due to cardiovascular causes and cardiovascular death. Secondary outcomes were histological atherosclerotic carotid plaque characteristics.

### Genotyping

Methods for genotyping, quality control and imputation in the Athero-Express biobank have been published elsewhere (25, 26). Briefly, DNA was extracted from EDTA whole blood samples or if not present from atherosclerotic plaque tissue according to validated protocols. Genotyping was performed with two commercially available chips: the first batch by Affymetrix Genome-Wide Human SNP array 5.0 (previously used in the Athero-Express Genomics Study 1 (AEGS1), covering samples obtained in 2002-2007) and the second batch by Affymetrix Axiom GW CEU 1 array (previously used in Athero-Express Genomics Study 2 (AEGS2), covering samples obtained in 2002-2013). Procedures for data quality control and data cleaning were in accordance with global standards.(27) After genotype calling according to Affymetrix’ specification, data was filtered on 1) individual call rate > 97%, 2) genotype call rate > 97%, 3) minor allele frequencies (MAF) > 3%, 4) average heterozygosity rate ± 3.0 standard deviations, 5) relatedness (pi-hat > 0.20), 6) Hardy–Weinberg Equilibrium (HWE p < 1.0×10^−6^), and 7) population stratification (based on HapMap 2, release 22, b36) by excluding samples deviating more than 6 standard deviations from the average in 5 iterations during principal component analysis and by visual inspection.(25) After pre-phasing using SHAPEIT2 v2.644, a combined dataset of 1000 Genome (phase 3, version 5) and The Genome of the Netherlands Project release 5 was used as a reference for imputation with IMPUTE2 v2.3.0.(28)

### Polygenic risk score (MetaGRS)

To estimate the polygenic cardiovascular disease susceptibility for included patients in our cohort, we used the previously published polygenic risk score for CAD (MetaGRS).(19) Its construction was described elsewhere.(19) Briefly, the MetaGRS comprises 1.7 million genetic variants associated with CAD and was constructed through meta-analysis of three genomic risk scores: GRS46K (comprising 46,000 cardiometabolic genetic variants), FDR202 (including 202 genetic variants associated with CAD at false discovery rate p<0.05 in the recent GWAS CARDIoGRAMplusC4D), and the 1000Genomes genetic score also created with CARDIoGRAMplusC4D. The MetaGRS was internally and externally validated for the primary risk of prevalent and incident CAD in the UK Biobank.(19) We calculated the MetaGRS for each included patient in this study and standardized it to mean-zero and unit-variance for each genotyping batch separately, i.e. AEGS1 and AEGS2, respectively.

### Sample handling

After CEA, the atherosclerotic plaque was directly processed in the laboratory following standardized protocols.(4, 23, 29, 30) The plaque was cut in cross-sectional segments of 5mm. The segment with largest plaque burden was chosen as the culprit lesion and immunohistochemically analysed for macrophages, smooth muscle cells (SMC), lipid core, calcification, collagen, intraplaque haemorrhage (IPH) and microvessel content. Extensive description of the standardized protocol for atherosclerotic plaque processing and analysis of plaque characteristics has been previously reported and is added to the Supplemental. (4, 23, 29, 30) To assess the overall vulnerability of the atherosclerotic plaque, a vulnerability score was created ranging from 0-5 with 1 point for plaque characteristics that are considered hallmarks of a vulnerable plaque (moderate/heavy macrophages, no/minor collagen, no/minor SMC, lipid core>10% and presence of IPH), based on a previous publication.(31)

### Statistical analysis

Baseline characteristics were compared between patient groups (FHx and MetaGRS) by chi-square test for categorical variables and Student’s t-test for continuous variables (lipid levels were log-transformed). We analysed the association between FHx and MetaGRS and sCVE by Cox-proportional hazard regression models and the associations with plaque characteristics through logistic or linear regression models. To fully unravel the genetic association, the association of MetaGRS with sCVE and plaque characteristics was analysed in three ways: (1) MetaGRS as a continuous quantification of genetic CAD susceptibility (2) patients in the top 20% of the MetaGRS distribution compared to the remaining 80%, and (3) patients in the top 20% of the MetaGRS distribution compared to those in the bottom 20% of the distribution. Kaplan-Meier curves were constructed to graphically illustrate univariate associations. Confounders for multivariable analyses were selected based on literature(19, 31, 32) (for sCVE these were age, sex, diabetes, BMI, smoking and hypercholesterolemia and for plaque characteristics these were age, sex, surgery year and type of cerebrovascular symptoms). Additional confounders were added when showing an association of p<0.20 with the determinant (FHx or MetaGRS) and outcome of interest (sCVE or plaque characteristics). For MetaGRS models genotype array and principal components 1-4 were also added. Full model description is displayed in the Supplemental Tables S1 and S2. Because a previous study in our biobank showed that IPH is associated with sCVE(4), IPH was added to multivariable models of FHx, MetaGRS and sCVE to explore whether IPH could be one possible underlying mechanism. Sex-stratified analyses were performed to unravel sex-dependent differences in associations. Values with p<0.05 were considered statistically significant. All analyses were performed in IBM SPSS Statistics version 25.0.

## Results

Patient selection from the Athero-Express Biobank and characteristics of the study population are displayed in Figure 1 and Table 1. Patients had a mean age 69 years and 70% were men. The cohort represented a typically severe atherosclerotic cohort with high prevalence of traditional risk factors and atherosclerotic manifestations in other vascular beds (coronary or peripheral arteries, respectively 30% and 20%). Baseline characteristics were similar between the total cohort with FHx data and the sub-cohort with genotyped data (Table 1).

**Table 1.**
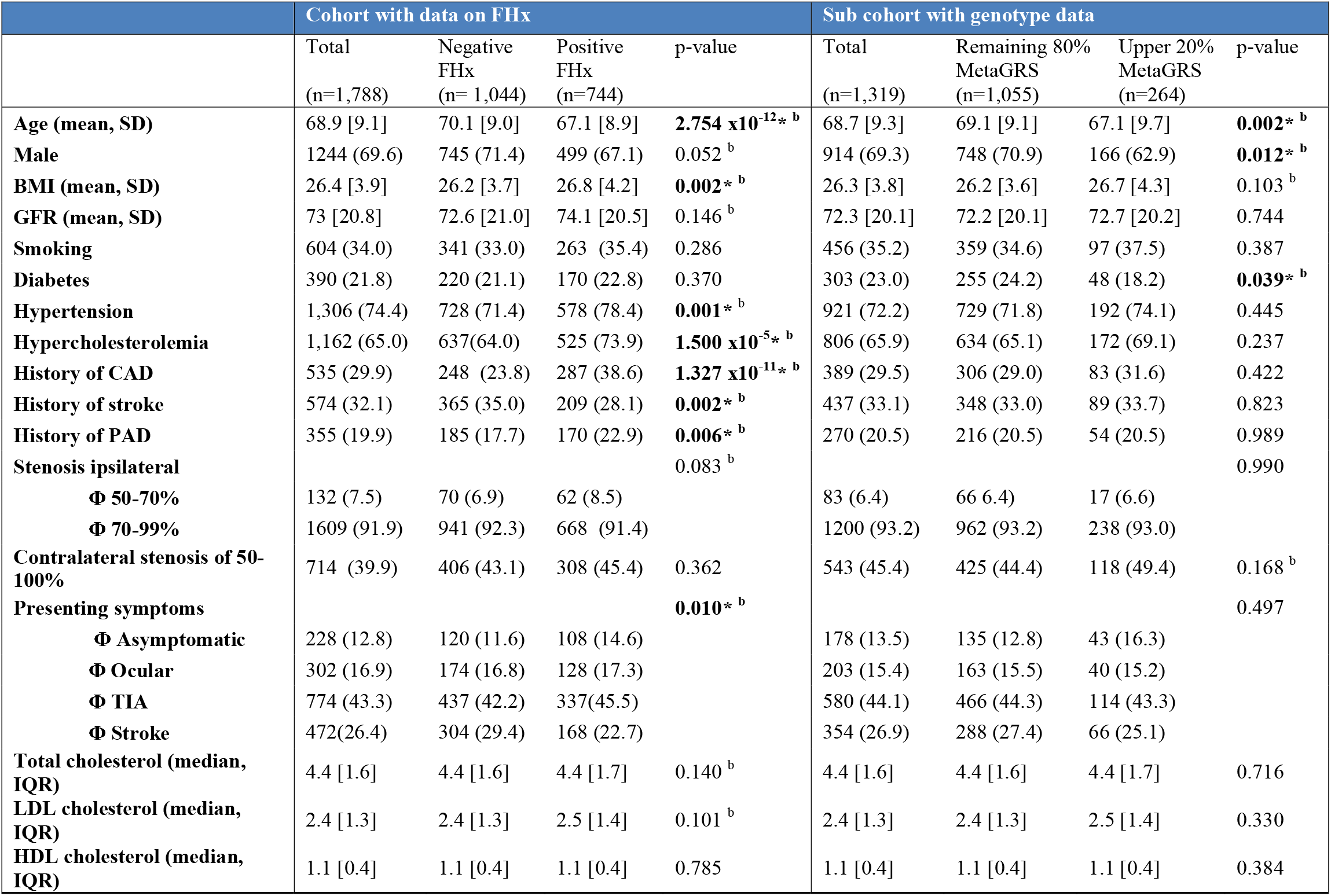
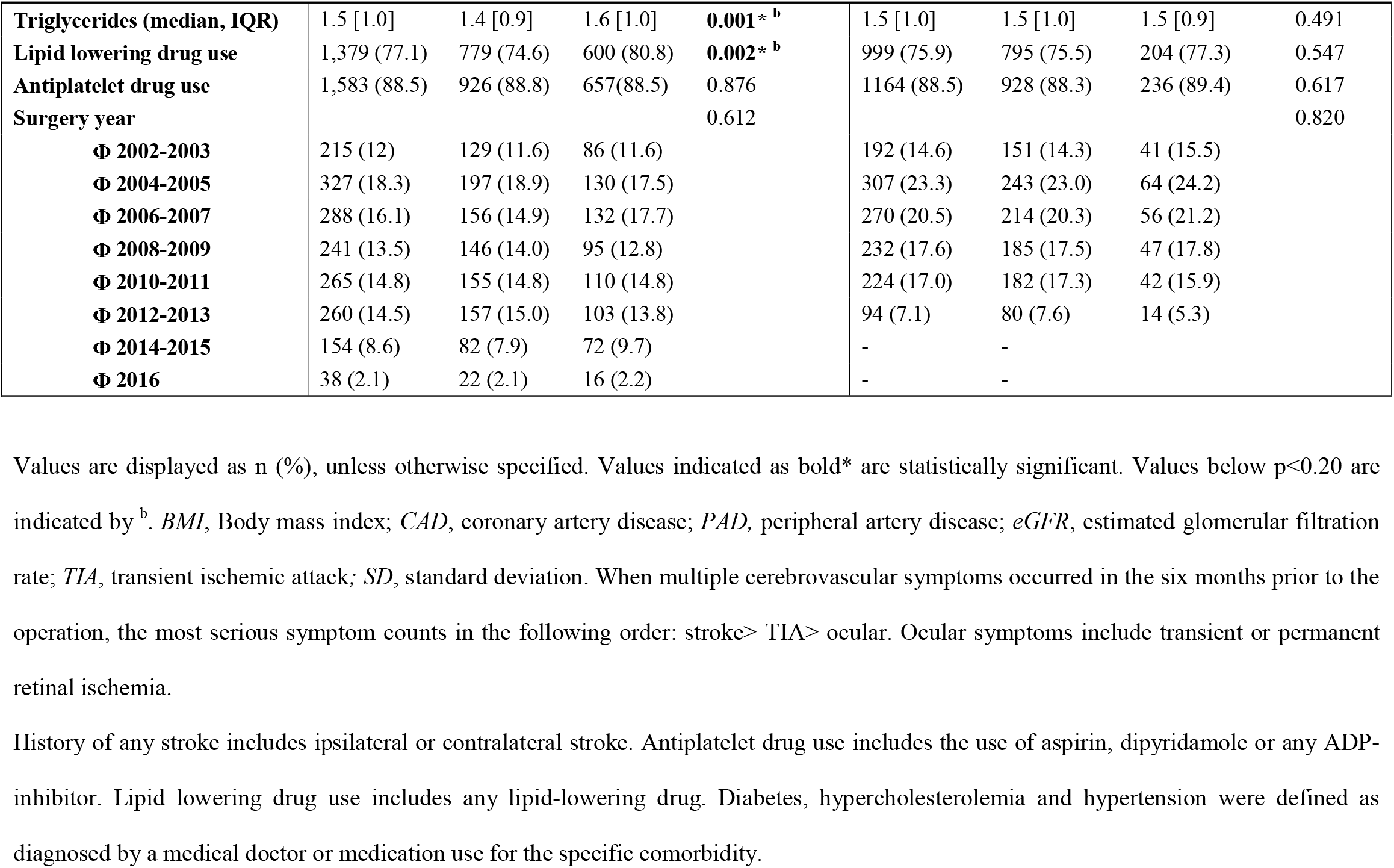
Baseline characteristics.

|  | Cohort with data on FHx |  |  |  | Sub cohort with genotype data |  |  |  |
| --- | --- | --- | --- | --- | --- | --- | --- | --- |
|  | Total<br>(n=1,788) | Negative<br>FHx<br>(n= 1,044) | Positive<br>FHx<br>(n=744) | p-value | Total<br>(n=1,319) | Remaining 80%<br>MetaGRS<br>(n=1,055) | Upper 20%<br>MetaGRS<br>(n=264) | p-value |
| **Age (mean, SD)** | 68.9 [9.1] | 70.1 [9.0] | 67.1 [8.9] | **2.754 x10<sup>-12</sup>\*<sup>b</sup>** | 68.7 [9.3] | 69.1 [9.1] | 67.1 [9.7] | **0.002\*<sup>b</sup>** |
| **Male** | 1244 (69.6) | 745 (71.4) | 499 (67.1) | 0.052<sup>b</sup> | 914 (69.3) | 748 (70.9) | 166 (62.9) | **0.012\*<sup>b</sup>** |
| **BMI (mean, SD)** | 26.4 [3.9] | 26.2 [3.7] | 26.8 [4.2] | **0.002\*<sup>b</sup>** | 26.3 [3.8] | 26.2 [3.6] | 26.7 [4.3] | 0.103<sup>b</sup> |
| **GFR (mean, SD)** | 73 [20.8] | 72.6 [21.0] | 74.1 [20.5] | 0.146<sup>b</sup> | 72.3 [20.1] | 72.2 [20.1] | 72.7 [20.2] | 0.744 |
| **Smoking** | 604 (34.0) | 341 (33.0) | 263 (35.4) | 0.286 | 456 (35.2) | 359 (34.6) | 97 (37.5) | 0.387 |
| **Diabetes** | 390 (21.8) | 220 (21.1) | 170 (22.8) | 0.370 | 303 (23.0) | 255 (24.2) | 48 (18.2) | **0.039\*<sup>b</sup>** |
| **Hypertension** | 1,306 (74.4) | 728 (71.4) | 578 (78.4) | **0.001\*<sup>b</sup>** | 921 (72.2) | 729 (71.8) | 192 (74.1) | 0.445 |
| **Hypercholesterolemia** | 1,162 (65.0) | 637(64.0) | 525 (73.9) | **1.500 x10<sup>-5</sup>\*<sup>b</sup>** | 806 (65.9) | 634 (65.1) | 172 (69.1) | 0.237 |
| **History of CAD** | 535 (29.9) | 248 (23.8) | 287 (38.6) | **1.327 x10<sup>-11</sup>\*<sup>b</sup>** | 389 (29.5) | 306 (29.0) | 83 (31.6) | 0.422 |
| **History of stroke** | 574 (32.1) | 365 (35.0) | 209 (28.1) | **0.002\*<sup>b</sup>** | 437 (33.1) | 348 (33.0) | 89 (33.7) | 0.823 |
| **History of PAD** | 355 (19.9) | 185 (17.7) | 170 (22.9) | **0.006\*<sup>b</sup>** | 270 (20.5) | 216 (20.5) | 54 (20.5) | 0.989 |
| **Stenosis ipsilateral** |  |  |  | 0.083<sup>b</sup> |  |  |  | 0.990 |
| **Φ 50-70%** | 132 (7.5) | 70 (6.9) | 62 (8.5) |  | 83 (6.4) | 66 6.4) | 17 (6.6) |  |
| **Φ 70-99%** | 1609 (91.9) | 941 (92.3) | 668 (91.4) |  | 1200 (93.2) | 962 (93.2) | 238 (93.0) |  |
| **Contralateral stenosis of 50-100%** | 714 (39.9) | 406 (43.1) | 308 (45.4) | 0.362 | 543 (45.4) | 425 (44.4) | 118 (49.4) | 0.168<sup>b</sup> |
| **Presenting symptoms** |  |  |  | **0.010\*<sup>b</sup>** |  |  |  | 0.497 |
| **Φ Asymptomatic** | 228 (12.8) | 120 (11.6) | 108 (14.6) |  | 178 (13.5) | 135 (12.8) | 43 (16.3) |  |
| **Φ Ocular** | 302 (16.9) | 174 (16.8) | 128 (17.3) |  | 203 (15.4) | 163 (15.5) | 40 (15.2) |  |
| **Φ TIA** | 774 (43.3) | 437 (42.2) | 337(45.5) |  | 580 (44.1) | 466 (44.3) | 114 (43.3) |  |
| **Φ Stroke** | 472(26.4) | 304 (29.4) | 168 (22.7) |  | 354 (26.9) | 288 (27.4) | 66 (25.1) |  |
| **Total cholesterol (median, IQR)** | 4.4 [1.6] | 4.4 [1.6] | 4.4 [1.7] | 0.140<sup>b</sup> | 4.4 [1.6] | 4.4 [1.6] | 4.4 [1.7] | 0.716 |
| **LDL cholesterol (median, IQR)** | 2.4 [1.3] | 2.4 [1.3] | 2.5 [1.4] | 0.101<sup>b</sup> | 2.4 [1.3] | 2.4 [1.3] | 2.5 [1.4] | 0.330 |
| **HDL cholesterol (median, IQR)** | 1.1 [0.4] | 1.1 [0.4] | 1.1 [0.4] | 0.785 | 1.1 [0.4] | 1.1 [0.4] | 1.1 [0.4] | 0.384 |
| **Triglycerides (median, IQR)** | 1.5 [1.0] | 1.4 [0.9] | 1.6 [1.0] | **0.001\***<sup>b</sup> | 1.5 [1.0] | 1.5 [1.0] | 1.5 [0.9] | 0.491 |
| **Lipid lowering drug use** | 1,379 (77.1) | 779 (74.6) | 600 (80.8) | **0.002\***<sup>b</sup> | 999 (75.9) | 795 (75.5) | 204 (77.3) | 0.547 |
| **Antiplatelet drug use** | 1,583 (88.5) | 926 (88.8) | 657(88.5) | 0.876 | 1164 (88.5) | 928 (88.3) | 236 (89.4) | 0.617 |
| **Surgery year** |  |  |  | 0.612 |  |  |  | 0.820 |
| **Φ 2002-2003** | 215 (12) | 129 (11.6) | 86 (11.6) |  | 192 (14.6) | 151 (14.3) | 41 (15.5) |  |
| **Φ 2004-2005** | 327 (18.3) | 197 (18.9) | 130 (17.5) |  | 307 (23.3) | 243 (23.0) | 64 (24.2) |  |
| **Φ 2006-2007** | 288 (16.1) | 156 (14.9) | 132 (17.7) |  | 270 (20.5) | 214 (20.3) | 56 (21.2) |  |
| **Φ 2008-2009** | 241 (13.5) | 146 (14.0) | 95 (12.8) |  | 232 (17.6) | 185 (17.5) | 47 (17.8) |  |
| **Φ 2010-2011** | 265 (14.8) | 155 (14.8) | 110 (14.8) |  | 224 (17.0) | 182 (17.3) | 42 (15.9) |  |
| **Φ 2012-2013** | 260 (14.5) | 157 (15.0) | 103 (13.8) |  | 94 (7.1) | 80 (7.6) | 14 (5.3) |  |
| **Φ 2014-2015** | 154 (8.6) | 82 (7.9) | 72 (9.7) |  | - | - |  |  |
| **Φ 2016** | 38 (2.1) | 22 (2.1) | 16 (2.2) |  | - | - |  |  |

Values are displayed as n (%), unless otherwise specified. Values indicated as bold\* are statistically significant. Values below p<0.20 are indicated by <sup>b</sup>. *BMI*, Body mass index; *CAD*, coronary artery disease; *PAD*, peripheral artery disease; *eGFR*, estimated glomerular filtration rate; *TIA*, transient ischemic attack; *SD*, standard deviation. When multiple cerebrovascular symptoms occurred in the six months prior to the operation, the most serious symptom counts in the following order: stroke> TIA> ocular. Ocular symptoms include transient or permanent retinal ischemia.
History of any stroke includes ipsilateral or contralateral stroke. Antiplatelet drug use includes the use of aspirin, dipyridamole or any ADP-inhibitor. Lipid lowering drug use includes any lipid-lowering drug. Diabetes, hypercholesterolemia and hypertension were defined as diagnosed by a medical doctor or medication use for the specific comorbidity.

**Figure 1.**
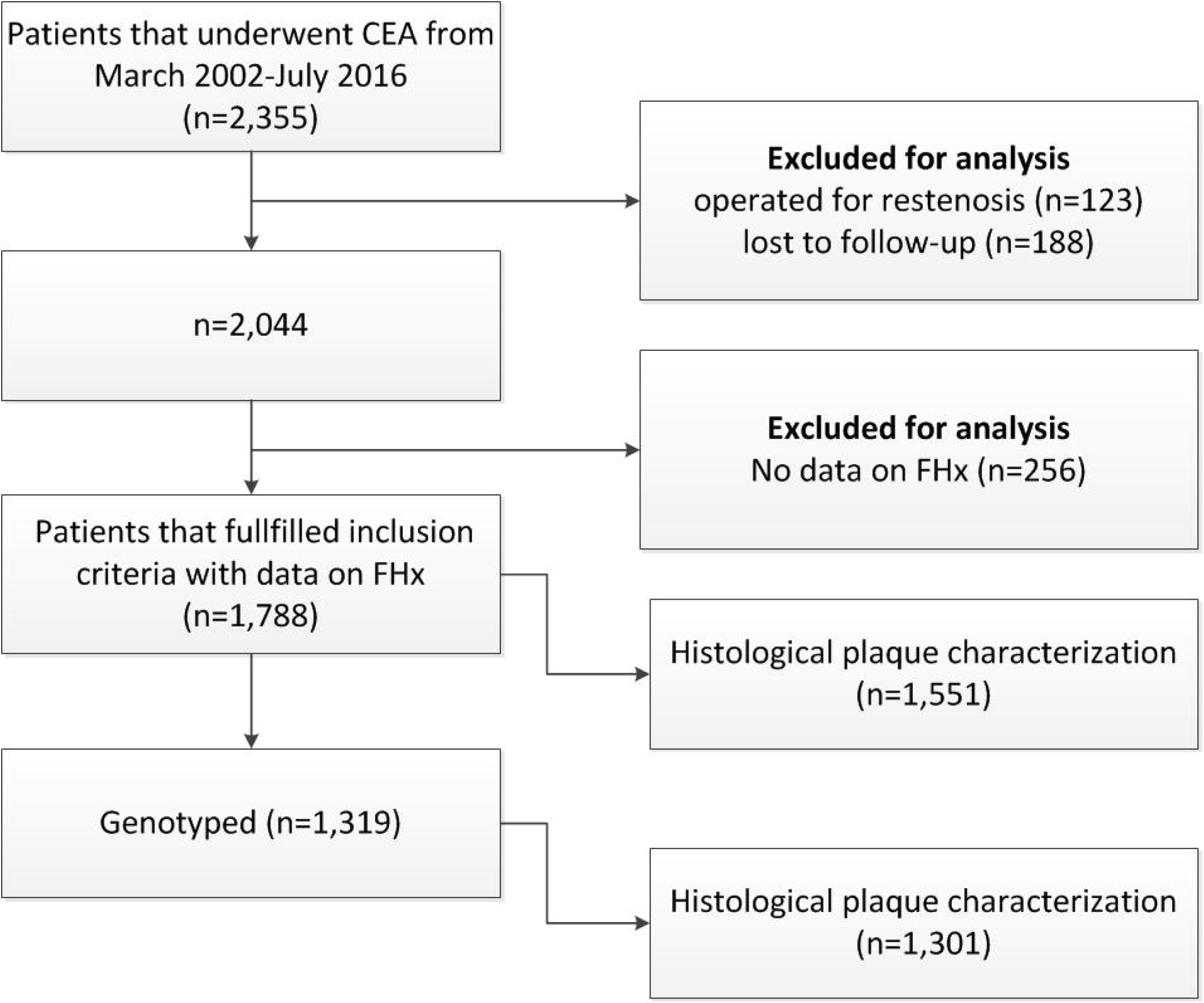
Flowchart.

### Patients with positive FHx have a higher risk of sCVE

Patients with a positive FHx (744/1,788, 41.6%) were younger and had on average more cardiovascular risk factors (Table 1). During a median follow-up of 2.9 years, 418 patients (23.4%) reached the composite endpoint of sCVE (Figure 2A) of whom 105 (5.9%) had stroke or fatal stroke, 119 (6.7%) had MI or fatal MI, 29 (1.6%) had cardiovascular death due to other causes (fatal cardiac failure, AAA rupture or sudden death) and 165 (9.2%) had a peripheral intervention or leg amputation.

**Figure 2.**
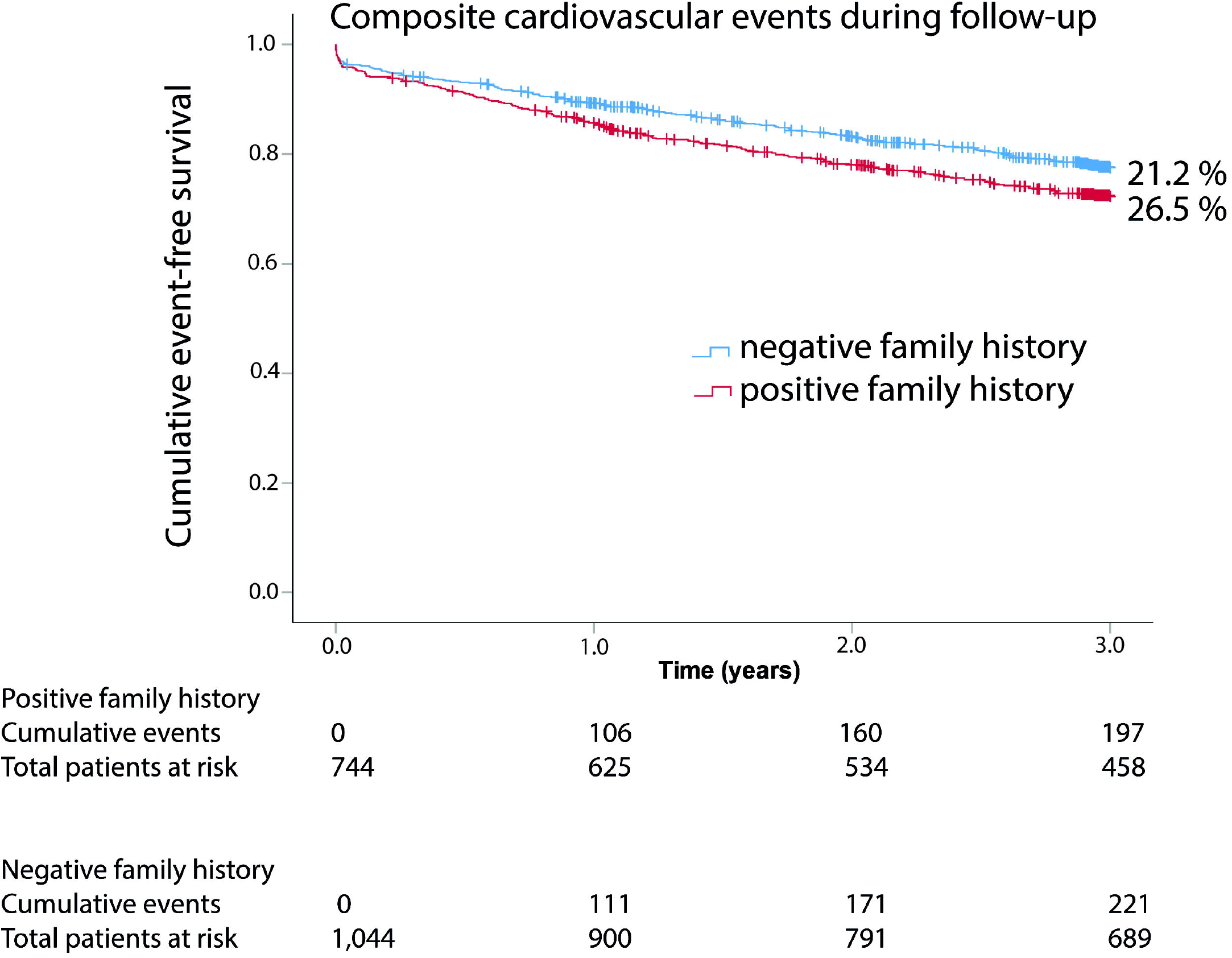
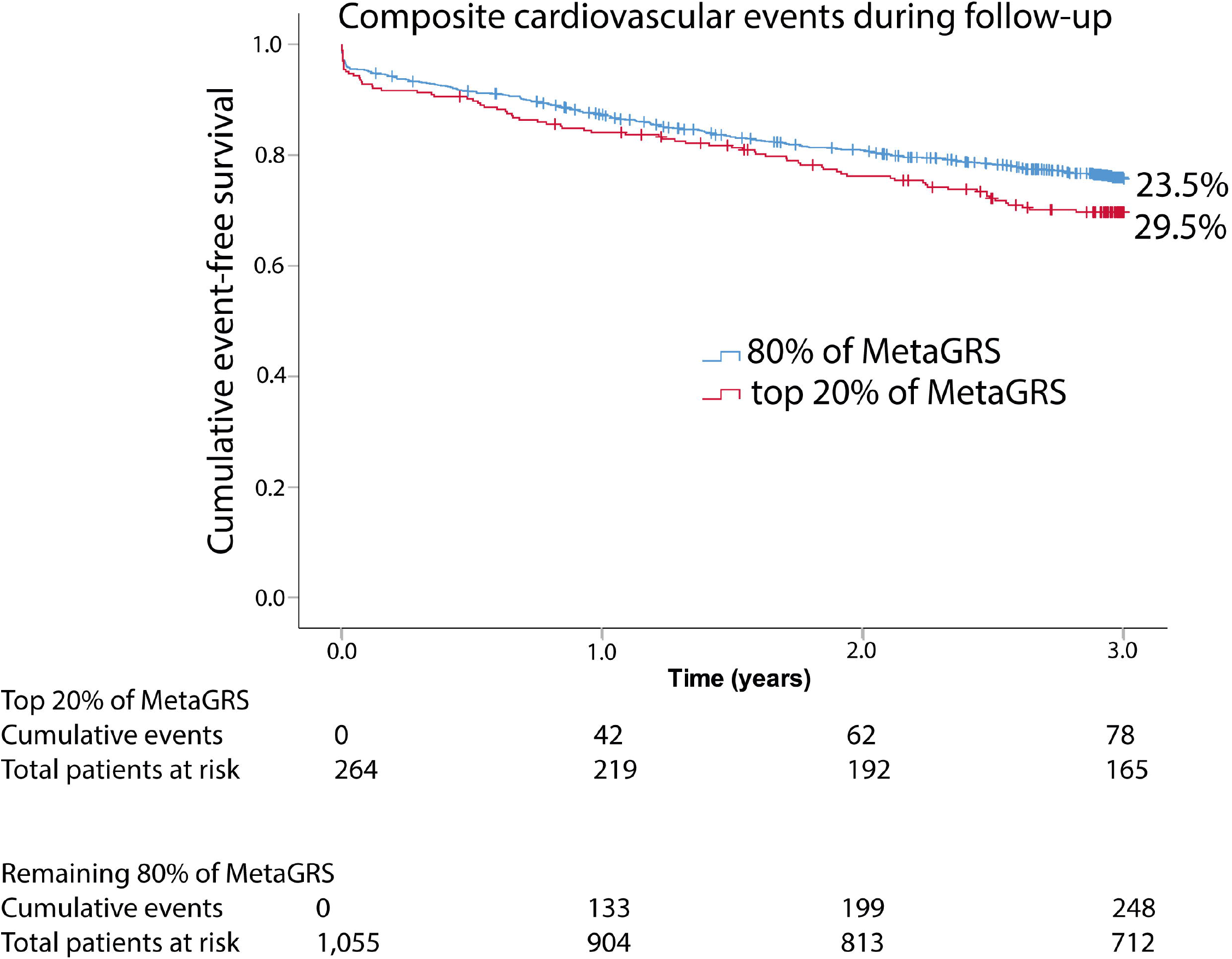
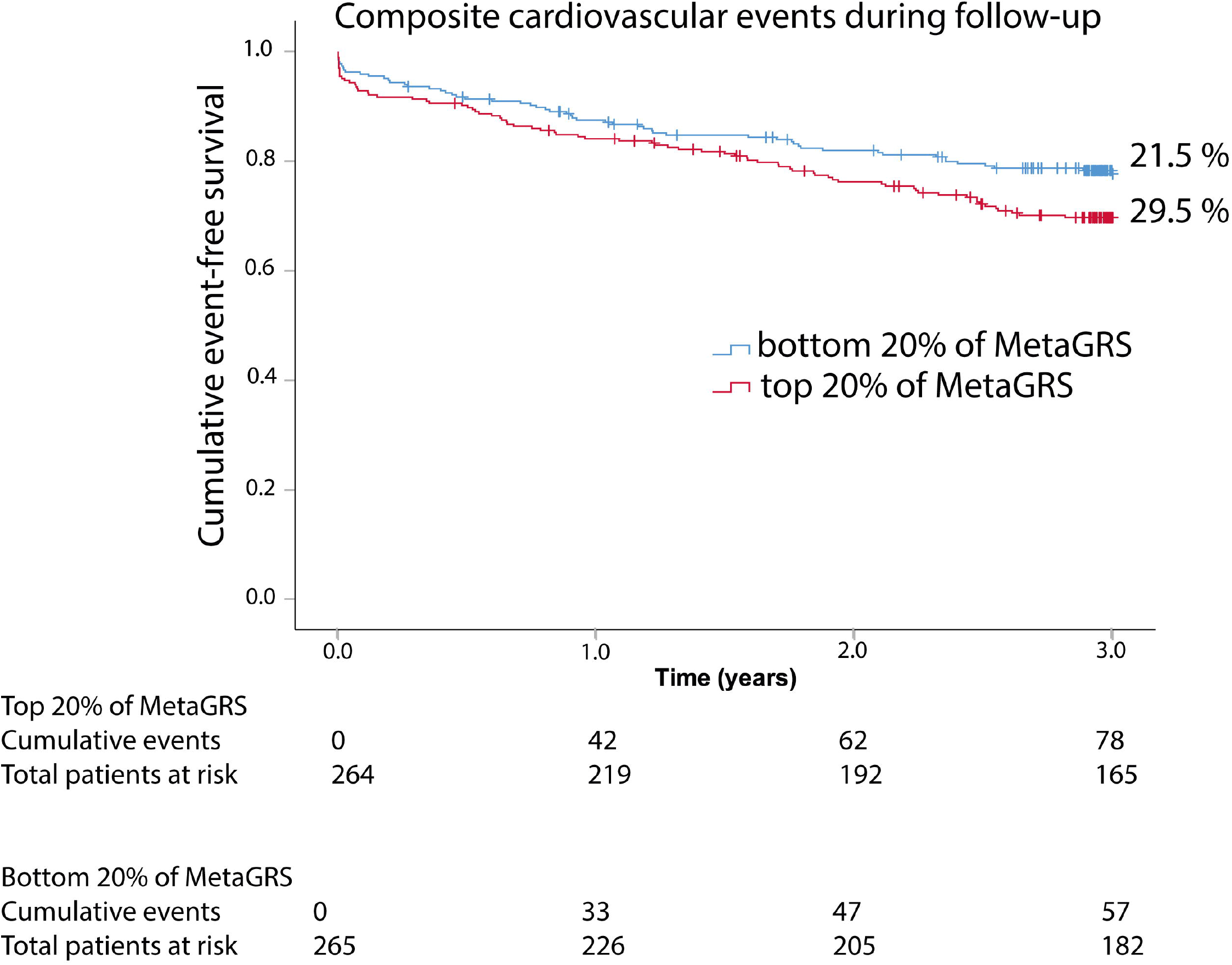
Kaplan-Meier graphs for 3-year risk of sCVE after CEA. 2A. Patients with positive FHx compared to those without. 2B. Patients in the top 20% of MetaGRS compared to the rest of the patients (remaining 80%). 2C. Patients in the top 20% of MetaGRS compared to those in the bottom 20% of MetaGRS.

Patients with positive FHx had an increased risk of sCVE compared to those without (absolute 3-year risks of 26.5% versus 21.2% respectively, hazard ratio (HR) 1.292, 95% confidence interval (CI), 1.066-1.566, p=0.009 (Figure 2A, Figure 3; Supplemental Table S3). This association remained significant after correction for confounders with adjusted HR 1.287, 95%CI 1.033-1.604, p=0.024 (Figure 3; Supplemental Table S3) and was independent of genetic predisposition as measured by MetaGRS (adjusted HR 1.397, 95%CI 1.074-1.819, p=0.013, Figure 3; Supplemental Table S3). Sex-stratified analyses confirmed results in men (with adjusted HR after correction for confounders of 1.380, 95%CI 1.068-1.783, p=0.014; adjusted HR after correction for confounders including MetaGRS was 1.513, 95%CI 1.115-2.052, p=0.008). However, in women the univariate association between FHx and sCVE was not significant (unadjusted HR 1.187, 95%CI 0.822-1.171, p=0.360) but multivariable analyses could not be performed because of limited power (Supplemental Table S3).

**Figure 3.**
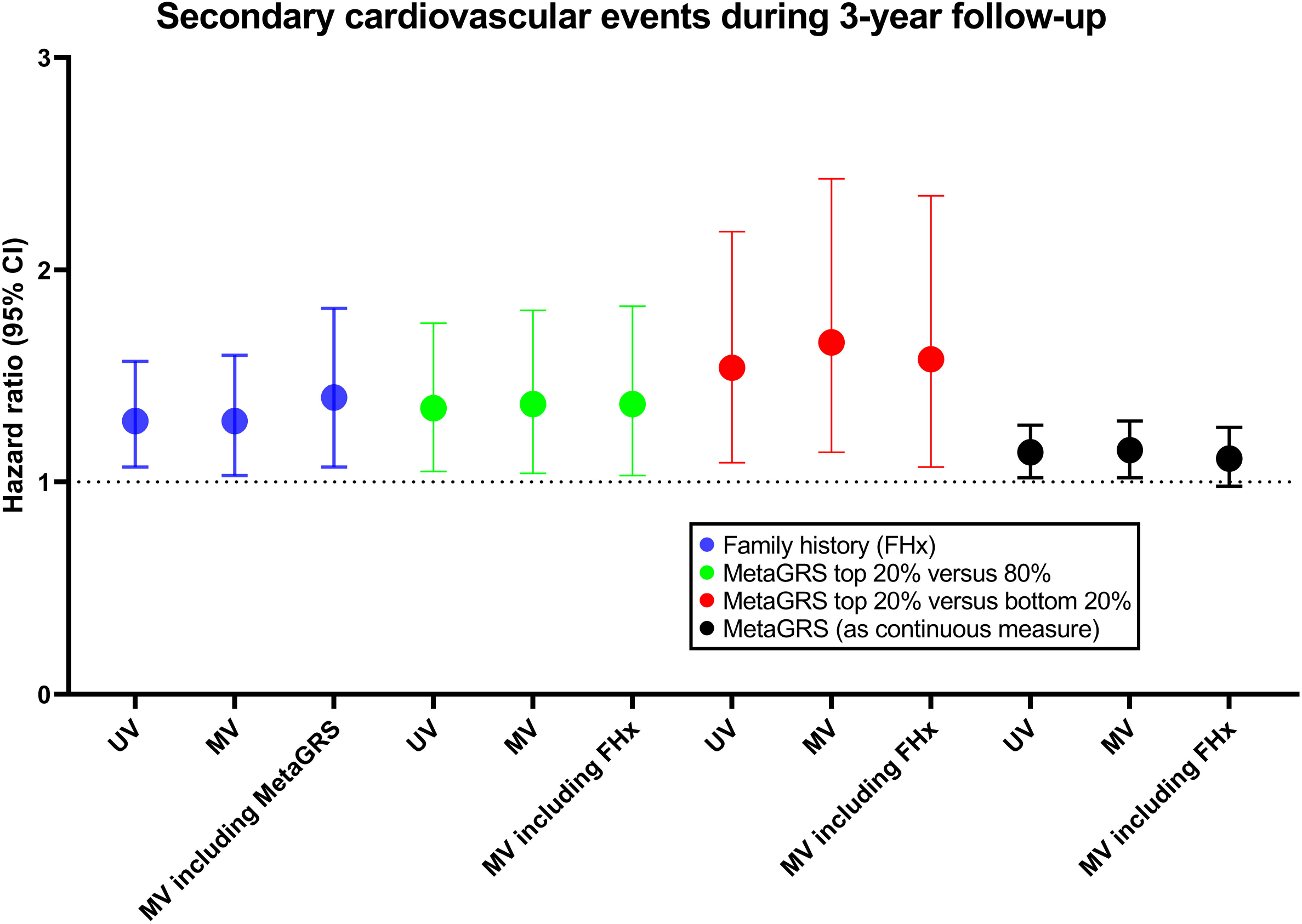
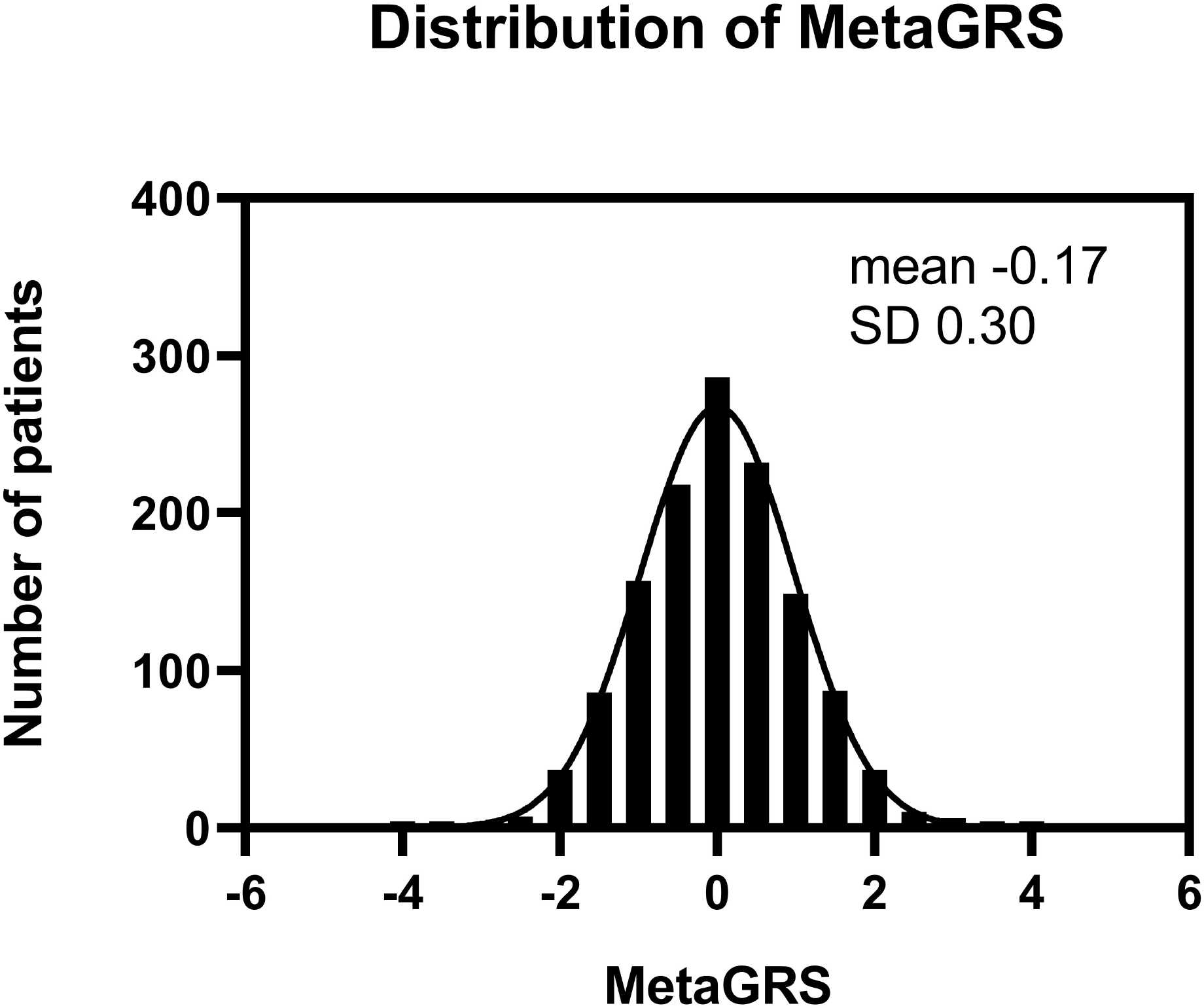
Cox-regression analyses of FHx and MetaGRs for sCVE after CEA. Hazard ratio’s (HR) and 95% confidence intervals (95% CI) for the different univariate and multivariable Cox-regression models of FHx or MetaGRS for sCVE.The HR for MetaGRS as a continuous quantity indicates the HR per one SD increase in MetaGRS. UV, univariate model. The univariate model for MetaGRS included age, sex, PC1-4 and genotype array. MV, multivariable model. The multivariable model for FHx was corrected for traditional risk factors (age, sex, hypercholesterolemia, diabetes, hypertension, BMI and smoking) and additional confounders (history of CAD, history of PAD, cerebrovascular symptoms and eGFR). The univariate model for MetaGRS included age, sex, PC1-4 and genotype array. For multivariable analyses of MetaGRS traditional risk factors (hypercholesterolemia, diabetes, hypertension, BMI and smoking) were added.

### Patients with higher MetaGRS have a higher risk of sCVE

The MetaGRS, standardized to mean-zero and unit-variance, approximated a normal distribution in the study population (Supplemental Figure S1). Patients in the top 20% of MetaGRS were relatively more often females, younger of age and had less often diabetes compared to the remaining 80% of the cohort (Table 1). Differences in baseline characteristics between high (highest quintile of MetaGRS) and low genetic risk patients (lowest quintile of MetGRS) can be found in the supplemental (Table S4). In the 3-year follow-up, a total of 326/1319 (24.7%) patients reached the composite endpoint of sCVE of whom 96 (7.3%) had stroke or fatal stroke, 85 (6.4%) had MI or fatal MI, 21 (1.6%) died of other cardiovascular causes (fatal cardiac failure, AAA rupture or sudden death) and 124 (9.4%) had a peripheral intervention or leg amputation.

Patients in the top 20% of MetaGRS had 1.4 fold increased risk of developing sCVE within the three years of follow-up when compared to the remaining 80% of the cohort (Figure 2B, absolute 3-year risks of 29.5% versus 23.5% respectively, HR 1.353, 95%CI 1.047-1.749, p=0.021). After adjustment for possible confounders including FHx this association remained statistically significant (HR for top 20% 1.345, 95%CI 1.009-1.792, p=0.043). We found similar results when we compared patients in top 20% to the bottom 20% of MetaGRS (in univariate analysis with HR 1.539, 95%CI,1.086-2.181, p=0.015 (Figure 2C) and for multivariable analysis adjusted

HR including FHx 1.583, 95%CI, 1.066-2.351, p=0.023), and when analysing MetaGRS as a continuous quantity (adjusted for confounders with HR 1.150 per one SD increase in MetaGRS, 95%CI 1.022-1.293, p=0.021, adjusted HR including FHx 1.112 per one SD increase in MetaGRS, 95% CI, 0.983-1.259, p=0.091). Results are illustrated in Figures 2B+C and 3, and displayed in Supplemental Table S3. Confounders added to multivariable models are displayed in Supplemental Tables S1 and S2.

Similar results were found in men (adjusted for confounders including FHx showed a HR 1.219 per one SD increase in MetaGRS, 95%CI 1.056-1.408, p=0.006, Supplemental Table S3). In women univariate analyses showed no significant associations between MetaGRS and sCVE (HR 0.916 per one SD increase in MetaGRS, 95% CI 0.743-1.129, p=0.413), yet multivariable analysis was not possible due to lack of power (Supplemental Table S3).

### MetaGRS is associated with vulnerable carotid plaque characteristics

To unravel possible underlying pathophysiological mechanisms of the associations between MetaGRS, FHx and CVD, we explored the impact of FHx and the MetaGRS on atherosclerotic plaque characteristics. We found no associations between histological plaque characteristics and FHx in the total cohort or in women although not all multivariable analyses could be performed (Supplemental Table S7 and S8). However carotid plaques from men with a positive FHx contained less collagen and less SMC content compared to men with negative FHx (Supplemental table S8). MetaGRS was associated with significantly higher overall plaque vulnerability score (regression coefficient β of 0.070 per SD increase in MetaGRS, 95%CI, 0.003-0.137, p=0.040). To determine the plaque characteristics on which this association was based, plaque characteristics were analysed separately. High genetic risk patients (in top 20% of MetaGRS) more frequently had a lipid core>10% of total plaque area (adjusted odds ratio (OR), 1.591, 95% CI 1.105-2.291, p=0.013) and more macrophage infiltration (adjusted OR 1.490, 95% CI 1.118-1.986, p=0.006) compared to patients with MetaGRS in the remaining 80% (Table 2). For lipid core, we found the same association when comparing the top 20% with the bottom 20% of the MetaGRS (adjusted HR 1.887, 95% CI, 1.188-2.997, p=0.007, Supplemental Table S5). Analyses of MetaGRS as a continuous quantity confirmed the association with lipid core>10% (adjusted OR 1.171 per SD increase in MetaGRS, 95% CI 1.026-1.337, p=0.019, Table 3). Sex-stratified analyses revealed a significant association of MetaGRS with macrophages in women (adjusted OR per SD increase in MetaGRS 1.238, 95%CI, 1.007-1.521, p=0.043) whereas a significant association of MetaGRS with IPH was found in men (adjusted OR per SD increase 1.220, 95%CI 1.050-1.418, p=0.010 Supplemental Table S6).

**Table 2.** Associations of carotid plaque characteristics from patients in top 20% of MetaGRS compared to the rest of the patients (80% of MetaGRS).

| Categorical plaque characteristics | 80% of<br>MetaGRS<br>(n=1,040) | Top 20% of<br>MetaGRS<br>(n=261) | Unadjusted OR<br>(95% CI) | Unadjusted<br>p-value | Adjusted OR<br>(95% CI) | Adjusted<br>p-value |
| --- | --- | --- | --- | --- | --- | --- |
| Moderate/heavy calcifications | 525/1,038 (50.6) | 135/261 (51.7) | 1.047 (0.798-1.374) | 0.741 | 1.001 (0.754-1.330) | 0.995 |
| Moderate/heavy collagen | 826/1,039 (79.5) | 211/261 (80.8) | 1.088 (0.772-1.534) | 0.629 | 1.091 (0.755-1.577) | 0.644 |
| Presence of lipid core >10% | 752/1,040 (72.3) | 209/261 (80.1) | 1.539 (1.104-2.147) | **0.011\*** | 1.591 (1.105-2.291) | **0.013\*** |
| Presence of IPH | 624/1,039 (60.1) | 160/261 (61.3) | 1.054 (0.798-1.392) | 0.713 | 1.112 (0.821-1.506) | 0.493 |
| Moderate/heavy smooth muscle cells | 736/1,037 (71.0) | 185/260 (71.2) | 1.009 (0.747-1.362) | 0.954 | 0.908 (0.652-1.265) | 0.570 |
| Moderate/heavy macrophages | 545/1,036 (52.6) | 161/260 (61.9) | 1.465 (1.109-1.936) | **0.007\*** | 1.490 (1.118-1.986) | **0.006\*** |
| Continuous plaque characteristics |  |  | Unadjusted beta<br>(95% CI) | Unadjusted<br>p-value | Adjusted beta<br>(95% CI) | Adjusted<br>p-value |
| Number of macrophages<br>(median, IQR) | 0.3 [1.0] | 0.5 [1.1] | 0.058 [-0.006-0.123] | 0.077 | 0.055 (-0.012-0.121) | 0.107 |
| Number of smooth muscle cells<br>(median, IQR) | 1.4 [2.3] | 1.3 [2.3] | -0.010 [-0.095-0.076] | 0.822 | -0.056 (-0.143-0.031) | 0.209 |
| Microvessels<br>(median, IQR) | 7.0 [7.0] | 7.3 [7.0] | 0.088 [-0.021-0.197] | 0.114 | 0.072 (-0.037-0.182) | 0.196 |
| Plaque vulnerability score | 2.0 [2.0] | 3 [1] | 0.216 (0.048-0.385) | **0.012\*** | 0.198 (0.003-0.364) | **0.004\*** |

**Table 3.** Associations of MetaGRS (as continuous measure) and histological carotid plaque characteristics.

| Categorical plaque characteristics | Unadjusted OR (95% CI) | Unadjusted p-value | Adjusted OR (95% CI) | Adjusted p-value |
| --- | --- | --- | --- | --- |
| Moderate/heavy calcifications | 0.977 (0.874-1.092) | 0.677 | 0.940 (0.826-1.070) | 0.348 |
| Moderate/heavy collagen | 1.035 (0.900-1.190) | 0.632 | 1.064 (0.919-1.231) | 0.408 |
| Presence of lipid core >10% | 1.148 (1.011-1.305) | **0.034\*** | 1.171 (1.026-1.337) | **0.019\*** |
| Presence of IPH | 1.101 (0.981-1.235) | 0.101 | 1.126 (0.999-1.270) | 0.052 |
| Moderate/heavy smooth muscle cells | 1.008 (0.889-1.143) | 0.899 | 1.004 (0.880-1.145) | 0.952 |
| Moderate/heavy macrophages | 1.095 (0.978-1.225) | 0.115 | 1.103 (0.983-1.237) | 0.095 |
| Continuous plaque characteristics | Unadjusted beta (95% CI) | Unadjusted p-value | Adjusted beta (95% CI) | Adjusted p-value |
| Number of macrophages (median, IQR) | 0.011 (-0.016-0.037) | 0.419 | 0.010 (-0.015-0.036) | 0.436 |
| Number of smooth muscle cells (median, IQR) | -0.007 (-0.041-0.027) | 0.647 | -0.004 (-0.038-0.031) | 0.837 |
| Microvessels (median, IQR) | 0.035 (-0.010-0.080) | 0.124 | 0.037 (-0.006-0.080) | 0.091 |
| Plaque vulnerability score | 0.072 (0.004-0.140) | **0.039\*** | 0.070 (0.003-0.137) | **0.040\*** |

Values are displayed as n/total (%), unless otherwise specified. Values indicated as bold\* are statistically significant.
MetaGRS was standardized to mean-zero and unit-variance. The odds ratio (OR) indicates OR per one SD increase in MetaGRS. *IPH*, intraplaque haemorrhage. Unadjusted OR indicates univariate analysis, adjusted OR indicates multivariable analysis. Multivariable analyses were corrected for age, sex, surgery year, cerebrovascular symptoms and additional confounders showing $p < 0.20$ association with MetaGRS and the plaque characteristics of interest.
Confounders included in various multivariable models can be found in Supplemental Table S2.

Because IPH has been associated with increased risk of sCVE(4), we added IPH to the multivariable models of FHx, MetaGRS and sCVE to unravel whether the association between FHx, MetaGRS and sCVE could be explained by IPH. We found that all associations of FHx and MetaGRS were independent of IPH given that adding IPH to multivariable models of sCVE did not alter the effect sizes (Supplemental Table S3).

## Discussion

We validated the polygenic risk score for CAD (MetaGRS) for sCVE in a severe atherosclerotic cohort of carotid artery stenosis patients undergoing carotid endarterectomy. We report four key findings. First, FHx was independently associated with an increased risk of sCVE. Second, higher MetaGRS score was independently associated with an increased risk of sCVE. Third, the association of MetaGRS with sCVE was independent of FHx and the association of FHx with sCVE was independent of MetaGRS. Fourth, high MetaGRS was associated with more vulnerable carotid atherosclerotic plaque characteristics suggesting possible underlying pathobiological mechanisms through which genetic variants could affect CVD.

Although positive FHx is a well-known risk factor for primary CVE(1, 2), previous studies assessing FHx and secondary outcome are inconsistent.(5, 7–9, 33) In patients with first-MI(7, 8, 33), studies have reported a protective effect of FHx on all-cause mortality, whereas others showed an increased risk of CVE.(5, 9) One can assume that patients with positive FHx are identified earlier as at-risk individuals through screening programs resulting in more intensive surveillance and preventive strategies leading to the benefit in overall survival.(7) Indeed, in our cohort patients with positive FHx were also younger at timing of CEA. Of note, it is known that the sensitivity of self-reported FHx can be low (50%-70%) and might therefore be an unreliable estimate.(34)

Our results are an independent validation of and consistent with a recent French Canadian study that revealed the association of MetaGRS in patients with recurrent CAD.(21) We now confirm this association in a different patient population with high prevalence of other CVD comorbidities either in coronary or peripheral vascular beds having severe carotid artery stenosis requiring intervention. Our results therefore underscore the concept of atherosclerosis as a complex and systemic disease underlying CVD. We extend the results from the French Canadian study as we corrected for FHx and possible confounding factors such as baseline medication, medical history and cardiovascular risk factors, highlighting the independent association of MetaGRS with sCVE. Furthermore, we provide mechanistic insights by showing associations of MetaGRS with vulnerable plaque characteristics (lipid core and macrophages content) indicative of an unstable overall plaque morphology. Previous studies in CAD patients assessing other CAD-GRS and sCVE were inconsistent, but these GRSs incorporated a limited number of SNPs (varying from 19 to 47 SNPs).(35–40) The current MetaGRS is the result of a meta-analysis and therefore captures information from the full genome for CAD, possibly explaining the clear reproducibility of this MetaGRS in two CAD cohorts and our CEA cohort.(19)

The independent association of FHx and PRS has been reported frequently for primary CAD events(19, 20, 41), however only one study showed this for sCVE.(40) We confirm the independent association of MetaGRS with sCVE from FHx. Indeed, we also found that positive FHx was associated with increased risk of sCVE independent of genetic risk (MetaGRS). Several reasons could be hypothesized for the non-overlapping associations of FHx and MetaGRS with CVE. MetaGRS includes common genetic variants associated to an increased risk for CAD in the general population, whilst CVD in families may arise in part from more rare genetic mutational events, and thus a positive FHx captures individual yet family specific rare variation. Another explanation could be that FHx reflects not only genetic factors but also non-genetic factors. Although we corrected for traditional risk factors, other environmental factors that were not taken into account such as social economic status or nutrition patterns could still be attributable to the risk of secondary events.

Exactly how genetic variants affect CVD is currently unknown. CAD-variants have been linked to pathways involved in atherosclerosis (e.g. lipid metabolism, inflammation, blood pressure and vascular remodelling), but most CAD-variants are situated outside protein-coding regions with unknown functions, making them hard to map to pathophysiological mechanisms.(13, 42) We found that MetaGRS was associated with a more rupture-prone atherosclerotic plaque displayed by a higher plaque vulnerability score caused by more fat, IPH (in men) and macrophages (predominantly in women). Moreover, carotid plaques from men with positive FHx were associated with less SMC and less collagen, whereas the association in women remains unclear. Similarly, a previous AE study showed that a CAD-PRS was correlated with more fat, whereas a large-artery stroke-PRS was correlated with more IPH and SMC.(18) Our results support the view that genetic variants could mediate their effect on CVD by influencing atherosclerotic plaque composition and morphology. Interestingly, IPH has been associated with sCVE in men independent of other clinical risk factors in our biobank (4, 43), yet we found that the associations of MetaGRS, FHx and sCVE were independent of IPH. It therefore remains unclear what the underlying mechanism is through which MetaGRS and FHx exert their increased sCVE risk. Moreover, it is shown that only small proportions (<8%) of the effect of genetic risk as well as of FHx for CAD were mediated through known metabolic pathways such as blood lipids and hypertension, whereas the majority (>80%) was not.(44) Albeit, these findings were based on a GRS consisting of only 50-CAD-SNPs. Future studies should explore exact pathophysiological mechanisms how genetic variants captured by the MetaGRS influence plaque destabilization, primary and secondary CVE risk, for example through deep-phenotyping of atherosclerotic plaque characteristics by quantitative computerized analysis(45), mapping MetaGRS loci to specific CVE (MI, stroke or PAD), and further investigating the potential sex-differences.

Although PRS could identify high-risk patients for CVE, one could conclude that due to their unfavourable genetic risk the CVE risk is unchangeable. However, previous studies have suggested that high genetic risk is modifiable by lifestyle interventions or medication, thus not deterministic per se.(46) Among individuals with high genetic risk (top 20% of CAD-GRS), those adhering to a healthy lifestyle had lower relative risk of a first-CAD event compared to those with an unfavourable lifestyle. (46) Moreover, both in primary or secondary prevention trials, patients with high genetic risk (those in top 20% of CAD-GRS) had a greater absolute and relative risk reduction of (recurrent) MI or coronary death than those with lower genetic risk despite equal LDL level reductions. (40, 47) In the current era of evolving new expensive drugs (e.g. PCSK9-inhibitors or new anti-inflammatory drugs such as IL1b-blockers(48)) and the increasing burden of CVD on the sustainability of the healthcare system, the cost-effectiveness ratio becomes important. In the future, it may be useful to investigate whether the efficacy of such medications varies across genetic risk categories for selecting those benefit most and qualify for these add-on therapies.

Admittedly, our study has several limitations. First, although our results may suggest interesting sex-differences as we only observed the independent association of MetaGRS and FHx for sCVE in men, the association in women remains unclear because we were underpowered for multivariable analysis. Second, we did not have data regarding medication use during follow-up nor therapy compliance. These factors could have interfered with observed sCVE rates. Second, owing to limited power we were unable to assess associations with separate CVE or determine the predictive value of MetaGRS above clinical risk factors. Last, most included patients are of European ancestries and generalizability to other ethnicities needs further attention. Yet, the Athero-Express Biobank is unique in its scope and its major strengths are 1) the unique population that is relatively unexplored in the field of FHx and PRS, 2) the prospective study design with detailed data on patient characteristics (including FHx), and 3) the extensive data on plaque composition and morphology that enable us to identify putative pathological mechanisms.

In the future, PRS may be a useful tool for personalized risk prediction for primary or secondary CVE. Adding MetaGRS to a model with traditional risk factors improved prediction of first-CAD events, although model improvement was modest.(19) The clinical utility of PRS for sCVE is still unknown because the incremental value of PRS above clinical factors still needs to be established. One study suggested an additive predictive value of PRS above clinical factors but did not include FHx (38), whereas other studies failed to demonstrate this.(35, 36, 39) The power of these studies may have been limited due to limited number of CVE. Pooling data of several cohorts including detailed data on preventive strategies and medications during follow-up together with use of uniform outcome definitions for sCVE and uniform PRS composition could help elucidate the clinical value of PRS, for example within international collaborations such as the GENIUS-CHD Consortium.(49) Furthermore, possible sex-differences in the role of risk prediction with PRS need to be further elucidated.

In conclusion, both higher MetaGRS and positive FHx were independently associated with increased risk of sCVE in carotid artery stenosis patients undergoing CEA. Higher MetaGRS was also associated with more vulnerable carotid plaque characteristics indicating possible underlying mechanism how genetic variants influence CVD. PRS could identify high-risk individuals and may help selecting future study populations when investigating new therapeutic CVD prevention strategies.

## Data Availability

Data will be available upon request.

## Acknowledgements

We thank Sara van Laar, Sara Bregman, Bram Vermeulen, Evelyn Velema and Joëlle van Bennekom for their technical support in the Athero-Express Biobank.

## Abbreviations

PRS: polygenic risk score
CVD: cardiovascular disease
CAD: coronary artery disease
SNP: single-nucleotide polymorphism
FHx: family history of premature CVD
MetaGRS: polygenic risk score for CAD including 1.7 million SNPs associated with CAD CEA carotid endarterectomy
CVE: cardiovascular event
sCVE: secondary cardiovascular event MI myocardial infarction
AAA: abdominal aortic aneurysm
ACS: acute coronary syndrome defined as a composite of myocardial infarction and unstable angina pectoris
PAD: peripheral artery disease
GWAS: large-scale genome wide association studies

## Conflict of interest statement

none

## Author contributions

Study design: NT, SH, SWvdL

Data collection: NT, SH, SWvdL, GP

Data analysis and interpretation: NT, SH, SWvdL

Drafting the article and figures: NT, SH, SWvdL

Critical revision and final approval: NT, SH, SWvdL, DPVdK, GJB, HMdR, FWA, GP

## Figure legends

**Central illustration. Family history and genetic risk score (MetaGRS) are independently associated with secondary cardiovascular events (sCVE) within 3-years after carotid endarterectomy (CEA)**.

In the figure the hazard ratio’s (HR) of the Cox-regression analyses are displayed for different multivariable models for the associaton with sCVE.

## Notes

Financial support, Dr. van der Laan was funded through grants from the Netherlands CardioVascular Research Initiative of the Netherlands Heart Foundation (CVON 2011/B019 and CVON 2017-20: Generating the best evidencebased pharmaceutical targets for atherosclerosis [GENIUS I&II]). Dr. Timmerman, Dr. De Borst, dr. De Kleijn, and dr. Pasterkamp are funded through the EU 755320 Taxinomisis grant. We acknowledge the European Research Area Network on Cardiovascular diseases (ERA-CVD, grant number 01KL1802). There are no competing interests declared.

### Competing Interest Statement

The authors have declared no competing interest.

### Funding Statement

Dr. van der Laan was funded through grants from the Netherlands CardioVascular Research Initiative of the Netherlands Heart Foundation (CVON 2011/B019 and CVON 2017-20: Generating the best evidence-based pharmaceutical targets for atherosclerosis [GENIUS I&II]). Dr. Timmerman, Dr. De Borst, dr. De Kleijn, and dr. Pasterkamp are funded through the EU 755320 Taxinomisis grant. We acknowledge the European Research Area Network on Cardiovascular diseases (ERA-CVD, grant number 01KL1802). There are no competing interests declared.

### Author Declarations

All relevant ethical guidelines have been followed and any necessary IRB and/or ethics committee approvals have been obtained.

Any clinical trials involved have been registered with an ICMJE-approved registry such as ClinicalTrials.gov and the trial ID is included in the manuscript.

## References

1. Lloyd-Jones DM, Nam B-H, D’Agostino, Sr RB, et al. Parental cardiovascular disease as a risk factor for cardiovascular disease in middle-aged adults. JAMA 2004;291:2204–2211.

2. Chow CK, Islam S, Bautista L, et al. Parental history and myocardial infarction risk across the world: The INTERHEART study. J Am Coll Cardiol 2011;57:619–27.

3. van den Berg MJ, Bhatt DL, Kappelle LJ, et al. Identification of vascular patients at very high risk for recurrent cardiovascular events: validation of the current ACC/AHA very high risk criteria. Eur Heart J 2017;38:3211–3218.

4. Hellings WE, Peeters W, Moll FL, et al. Composition of carotid atherosclerotic plaque is associated with cardiovascular outcome: a prognostic study. Circulation 2010;121:1941–50.

5. Mulders TA, Meyer Z, van der Donk C, et al. Patients with premature cardiovascular disease and a positive family history for cardiovascular disease are prone to recurrent events. Int J Cardiol 2011;153:64–67.

6. Canto JG, Kiefe CI, Rogers WJ, et al. Atherosclerotic risk factors and their association with hospital mortality among patients with first myocardial infarction (from the national registry of myocardial infarction). Am J Cardiol 2012;110:1256–1261.

7. Abdi-Ali A, Shaheen A, Southern D, et al. Relation between family history of premature coronary artery disease and the risk of death in patients with coronary artery disease. Am J Cardiol 2016;117:353–358.

8. Harpaz D, Behar S, Rozenman Y, Boyko V, Gottlieb S, Israeli Working Group on Intensive Cardiac Care IHS. Family history of coronary artery disease and prognosis after first acute myocardial infarction in a national survey. Cardiology 2004;102:140–146.

9. Kim C, Chang H-J, Cho I, et al. Impact of family history on the presentation and clinical outcomes of coronary heart disease: data from the Korea Acute Myocardial Infarction Registry. Korean J Intern Med 2013;28:547–56.

10. Nikpay M, Goel A, Won H-H et al. A comprehensive 1000 Genomes–based genome-wide association meta-analysis of coronary artery disease. Nat Genet 2015;47:1121–1130.

11. Howson JMM, Zhao W, Barnes DR, et al. Fifteen new risk loci for coronary artery disease highlight arterial-wall-specific mechanisms. Nat Genet 2017;49:1113–1119.

12. Verweij N, Eppinga RN, Hagemeijer Y, van der Harst P. Identification of 15 novel risk loci for coronary artery disease and genetic risk of recurrent events, atrial fibrillation and heart failure. Sci Rep 2017;7:2761.

13. Erdmann J, Kessler T, Munoz Venegas L, Schunkert H. A decade of genome-wide association studies for coronary artery disease: the challenges ahead. Cardiovasc Res 2018;114:1241–1257.

14. van der Harst P, Verweij N. Identification of 64 novel genetic loci provides an expanded view on the genetic architecture of coronary artery disease. Circ Res 2018;122:433–443.

15. Traylor M, Farrall M, Holliday EG, et al. Genetic risk factors for ischaemic stroke and its subtypes (the METASTROKE collaboration): a meta-analysis of genome-wide association studies. Lancet Neurol 2012;11:951–62.

16. Holliday EG, Maguire JM, Evans T-J, et al. Common variants at 6p21.1 are associated with large artery atherosclerotic stroke. Nat Genet 2012;44:1147–51.

17. Malik R, Traylor M, Pulit SL, e.a. Low-frequency and common genetic variation in ischemic stroke. Neurology 2016;86:1217–1226.

18. van der Laan SW, Siemelink MA, Haitjema S, et al. Genetic susceptibility loci for cardiovascular disease and their impact on atherosclerotic plaques. Circ Genomic Precis Med 2018;11:e002115.

19. Inouye M, Abraham G, Nelson CP, et al. Genomic risk prediction of coronary artery disease in 480,000 adults: implications for primary prevention. J Am Coll Cardiol 2018;72:1883–1893.

20. Khera AV, Chaffin M, Aragam KG, et al. Genome-wide polygenic scores for common diseases identify individuals with risk equivalent to monogenic mutations. Nat Genet 2018;50:1219–1224.

21. Wünnemann F, Sin Lo K, Langford-Avelar A, et al. Validation of genome-wide polygenic risk scores for coronary artery disease in french canadians. Circ Genomic Precis Med 2019;12:e002481.

22. Ntalla I, Kanoni S, Zeng L, et al. Genetic risk score for coronary disease identifies predispositions to cardiovascular and noncardiovascular diseases. J Am Coll Cardiol 2019;73:2932–2942.

23. Verhoeven BAN, Velema E, Schoneveld AH, et al. Athero-express: differential atherosclerotic plaque expression of mRNA and protein in relation to cardiovascular events and patient characteristics. Rationale and design. Eur J Epidemiol 2004;19:1127–33.

24. Kumar R, Batchelder A, Saratzis A, et al. Restenosis after carotid interventions and its relationship with recurrent ipsilateral stroke: a systematic review and meta-analysis. Eur J Vasc Endovasc Surg 2017;53:766–775.

25. van der Laan SW, Foroughi Asl H, van den Borne P, et al. Variants in ALOX5, ALOX5AP and LTA4H are not associated with atherosclerotic plaque phenotypes: The Athero-Express Genomics Study. Atherosclerosis 2015;239:528–538.

26. Siemelink MA, van der Laan SW, van Setten J, et al. Common variants associated with blood lipid levels do not affect carotid plaque composition. Atherosclerosis 2015;242:351–356.

27. Anderson CA, Pettersson FH, Clarke GM, Cardon LR, Morris AP, Zondervan KT. Data quality control in genetic case-control association studies. Nat Protoc 2010;5:1564–1573.

28. Genome of the Netherlands Consortium, Francioli LC, Menelaou A, et al. Whole-genome sequence variation, population structure and demographic history of the Dutch population. Nat Genet 2014;46:818–825.

29. Vrijenhoek JEP, Nelissen BGL, Velema E, et al. High reproducibility of histological characterization by whole virtual slide quantification; an example using carotid plaque specimens. PLoS One 2014;9:e115907.

30. Hellings WE, Pasterkamp G, Vollebregt A, et al. Intraobserver and interobserver variability and spatial differences in histologic examination of carotid endarterectomy specimens. J Vasc Surg 2007;46:1147–1154.

31. Verhoeven B, Hellings WE, Moll FL, et al. Carotid atherosclerotic plaques in patients with transient ischemic attacks and stroke have unstable characteristics compared with plaques in asymptomatic and amaurosis fugax patients. J Vasc Surg 2005;42:1075–1081.

32. van Lammeren GW, den Ruijter HM, Vrijenhoek JEP, et al. Time-dependent changes in atherosclerotic plaque composition in patients undergoing carotid surgery. Circulation 2014;129:2269–2276.

33. Canto JG, Kiefe CI, Rogers WJ, et al. Number of coronary heart disease risk factors and mortality in patients with first myocardial infarction. JAMA 2011;306:2120–2127.

34. Murabito JM, Nam B-H, D’Agostino RB, Lloyd-Jones DM, O’Donnell CJ, Wilson PWF. Accuracy of offspring reports of parental cardiovascular disease history: the framingham offspring study. Ann Intern Med 2004;140:434–40.

35. Weijmans M, de Bakker PIW, van der Graaf Y, et al. Incremental value of a genetic risk score for the prediction of new vascular events in patients with clinically manifest vascular disease. Atherosclerosis 2015;239:451–458.

36. Labos C, Martinez SC, Leo Wang RH, et al. Utility of a genetic risk score to predict recurrent cardiovascular events 1 year after an acute coronary syndrome: A pooled analysis of the RISCA, PRAXY, and TRIUMPH cohorts. Atherosclerosis 2015;242:261–267.

37. Christiansen MK, Nyegaard M, Larsen SB, et al. A genetic risk score predicts cardiovascular events in patients with stable coronary artery disease. Int J Cardiol 2017;241:411–416.

38. Wirtwein M, Melander O, Sjőgren M, et al. Relationship between selected DNA polymorphisms and coronary artery disease complications. Int J Cardiol 2017;228:814–820.

39. Vaara S, Tikkanen E, Parkkonen O, et al. Genetic risk scores predict recurrence of acute coronary syndrome. Circ Cardiovasc Genet 2016;9:172–178.

40. Mega JL, Stitziel NO, Smith JG, et al. Genetic risk, coronary heart disease events, and the clinical benefit of statin therapy: an analysis of primary and secondary prevention trials. Lancet 2015;385:2264–2271.

41. Tada H, Melander O, Louie JZ, et al. Risk prediction by genetic risk scores for coronary heart disease is independent of self-reported family history. Eur Heart J 2016;37:561–567.

42. Haitjema S, Meddens CA, van der Laan SW, et al. Additional candidate genes for human atherosclerotic disease identified through annotation based on chromatin organization. Circ Cardiovasc Genet 2017;10:e001664.

43. Vrijenhoek JEP, Den Ruijter HM, De Borst GJ, et al. Sex is associated with the presence of atherosclerotic plaque hemorrhage and modifies the relation between plaque hemorrhage and cardiovascular outcome. Stroke 2013;44:3318–23.

44. Fritz J, Shiffman D, Melander O, Tada H, Ulmer H. Metabolic mediators of the effects of family history and genetic risk score on coronary heart disease—findings from the Malmö Diet and Cancer Study. J Am Heart Assoc 2017;6:e005254.

45. Nelissen BGL, van Herwaarden JA, Moll FL, van Diest PJ, Pasterkamp G. SlideToolkit: an assistive toolset for the histological quantification of whole slide images. PLoS One 2014;9:e110289.

46. Khera AV, Emdin CA, Drake I, et al. Genetic risk, adherence to a healthy lifestyle, and coronary disease. N Engl J Med 2016;375:2349–2358.

47. Natarajan P, Young R, Stitziel NO, et al. Polygenic risk score identifies subgroup with higher burden of atherosclerosis and greater relative benefit from statin therapy in the primary prevention setting. Circulation 2017;135:2091–2101.

48. Ridker PM, Everett BM, Thuren T, et al. Antiinflammatory therapy with canakinumab for atherosclerotic disease. N Engl J Med. 2017;377:1119–1131.

49. Patel RS, Asselbergs FW. The GENIUS-CHD consortium. Eur Heart J 2015;36:2674–6.

